# MaSk-LMM: A Matrix Sketching Framework for Linear Mixed Models in Association Studies

**DOI:** 10.1101/2023.11.13.23298469

**Authors:** Myson Burch, Aritra Bose, Gregory Dexter, Laxmi Parida, Petros Drineas

**Affiliations:** IBM T.J. Watson Research Center, Yorktown Heights, NY; Department of Computer Science, Purdue University, West Lafayette, IN

**Keywords:** Linear Mixed Models, Matrix Sketching, GWAS

## Abstract

Linear mixed models (LMMs) have been widely used in genome-wide association studies (GWAS) to control for population stratification and cryptic relatedness. Unfortunately, estimating LMM parameters is computationally expensive, necessitating large-scale matrix operations to build the genetic relatedness matrix (GRM). Over the past 25 years, Randomized Linear Algebra has provided alternative approaches to such matrix operations by leveraging *matrix sketching*, which often results in provably accurate fast and efficient approximations. We leverage *matrix sketching* to develop a fast and efficient LMM method called **Ma**trix-**Sk**etching **LMM** (**MaSk-LMM**) by sketching the genotype matrix to reduce its dimensions and speed up computations. Our framework comes with both theoretical guarantees and a strong empirical performance compared to current state-of-the-art.

## 1 Introduction

Linear Mixed Models (LMMs) are widely used when conducting genome-wide association studies (GWAS) for quantitative traits in the presence of population structure. It is well-known that population structure plays an important role in confounding results and generating false positive associations [34]. LMMs are able to capture and correct such confounders in the data, while decomposing phenotypic correlations into genetic and non-genetic components. These desirable properties have resulted in wide use of LMMs in GWAS and genomic selection problems in human and plant genetics, as well as in other biological applications [17, 22, 23, 31, 32].

On the negative side, LMMs have well-known limitations that we attempt to address in our work. Most promiment among those limitations are the increased computational requirements in terms of computatational time and memory space that these models necessitate. Computing LMM parameters involves building a genetic relationship matrix (GRM) to account for genome-wide sample structure; estimating the phenotypic variance using a random-effects model; and computing association statistics that account for the variance. LMMs require multiple 𝒪(*n*^3^) or 𝒪(*mn*^2^) matrix operations such as large matrix inversions, multiplications, etc. (Here *m* is the number of Single Nucleotide Polymorphisms (SNPs) or genetic markers and *n* is the number of individual samples in the study.) Such operations make straight-forward LMM computations intractable for large biobanks and create a need for methods that reduce the computational cost of LMM association analyses. Several methods have been developed to achieve computational speedups: Prominent among those are EMMAX [15], FaST-LMM [17], GEMMA [36], GRAMMAR-Gamma [25], GCTA [32], BOLT-LMM [18], Regenie [19], FastGWA [13], and SAIGE [35]. Some of these methods compute the LMM variance parameter exactly and obtain speedups using spectral decompositions of the GRM [15] via block optimizations [17]. Other methods perform approximate variance estimation [15, 25], while BOLT-LMM, fastGWA, Regenie, and SAIGE all perform a two-step procedure, where in the first step a model is fitted to a smaller set of genome-wide markers and in the second step a larger set of imputed variants are tested for association using the model estimates from the first step [20].

To the best of our knowledge, while prior work has been widely successful in significantly reducing the running time of LMMs in biobank-scale datasets by using optimized implementations and heuristic approaches, there is an alarming lack of theoretical underpinnings of such methods that could provide insights on the accuracy of the heuristics that have been used to speed up LMM computations. Additionally, recent advances in applied mathematics leveraging *matrix sketching* to speed up matrix computations (such as matrix inversion, log-determinant computations, etc.) that are major computational bottlenecks for LMMs have not been systematically explored, either theoretically or empirically, in prior work.

We propose and evaluate a method based on *Matrix-Sketching LMM (MaSk-LMM)*, to approximately solve LMMs by applying sketching to the original genotype matrix to reduce both its dimensions, while preserving the relevant properties of the original matrix for LMM computations. We provide theoretical support to our sketching approach by proving (see Theorem 3 in Appendix) that sketching the genetic markers (columns) of the genotype matrix results in bounded accuracy loss for the underlying LMM. To the best of our knowledge, this is the first theoretical result of its type, arguing that dimensionality reduction on the genetic marker space (which is typically massive in modern genetic datasets) is feasible without a significant loss in accuracy. Beyond our theoretical guarantees, we demonstrate that using simulated data and solving the LMM using the sketched matrix yields a similar number of causal and spurious genetic associations when compared to the solution using the original matrix. When applied to data for complex diseases, we recover previously known associations as well as novel loss of function (LoF) markers, which are possibly associated with coronary artery disease and hypertension. In both synthetic and real data, we observe speed-ups using our approach compared to Regenie, BOLT-LMM and FaST-LMM.

## 2 Materials and Methods

### 2.1 Mixed-model association

Linear mixed models (LMMs) are formed using the following simple linear model:

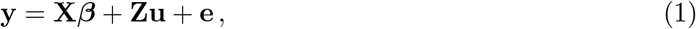

where^3^ **y** ∈ ℝ^*n*^ is the measured phenotype (response); **X** ∈ ℝ^*n×k*^ is the matrix of the *k* covariates (*e*.*g*. principal components, age, sex, etc.) with the corresponding vector of fixed effects *β* ∈ ℝ^*k*^; **Z** ∈ ℝ^*n×m*^ is the genotype matrix of *n* individuals genotyped on *m* genetic markers with **u** ∈ ℝ^*m*^ being the corresponding genetic effects vector; and **e** ∈ ℝ^*n*^ is the error vector or the component of **y** which cannot be explained by the model. We assume **u** and **e** are independent vectors and moreover that^4^ 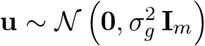 and 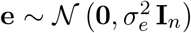 with scalars 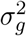 and 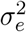 being the heritable and non-heritable components of **u** and **e** respectively. In the LMM setting, some form of maximum likelihood estimation is used to estimate the random and fixed effects of the model in order to identify genetic associations while correcting for confounding effects.

### 2.2 MaSk-LMM

Our approach, MaSk-LMM, mitigates the computational complexity of LMMs by using sample and marker sketching on the input genotype matrix **Z**, as well as on the response vector **y**. This allows us to significantly reduce the dimensions of the genotype matrix, as well as of the relatedness or kinship matrix (GRM). As discussed in the introduction, sketching reduces the dimensions of the input while maintaining sufficient information to approximate functions of the original input accurately. Let 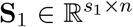 and 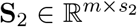 be two sketching matrices, with *s*_1_ ≪ *n* and *s*_2_ ≪ *m*. Here *s*_1_ and *s*_2_ are the sketching dimensions and are user-controlled parameters. Simple constructions for **S**_1_ and **S**_2_ are to have their entries drawn in independent identical trials from a Gaussian distribution of zero mean and variance 1*/s*_1_ and 1*/s*_2_, respectively. We can then use **S**_1_ and **S**_2_ to sketch the input genotype matrix as follows:

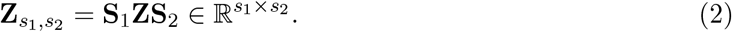

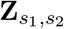 is computed in blocks so the entire original input does not need to be loaded into memory alleviating a portion of the computational burden of this approach. Notice that 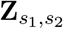 is now a much smaller *s*_1_ × *s*_2_ matrix which can be used in downstream computations instead of **Z**. For example, we can approximate the GRM as follows:

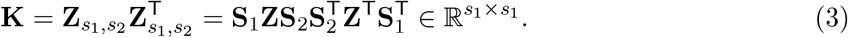

We also sketch the *n*-dimensional response vector **y** to construct the *s*_1_-dimensional response vector 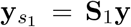 to be used in downstream computations instead of **y**. It is worth noting that there is a long line of research on matrix sketching methods, including gaussian sketching, the use of the subsampled randomized hadamard transforms, the count-min sketch, etc. and its application in human genetics [1–3]. In our work, we evaluated both the count-min sketch and the gaussian sketch. Both methods performed similarly and we chose to report results on gaussian sketching only, because it is conceptually simpler as well as easier to implement and theoretically analyze. See [29] for a discussion of other sketching methods and their theoretical properties. Figure 1 summarizes our framework and Algorithm 1 provides a high-level overview of our approach.

**Fig. 1:**
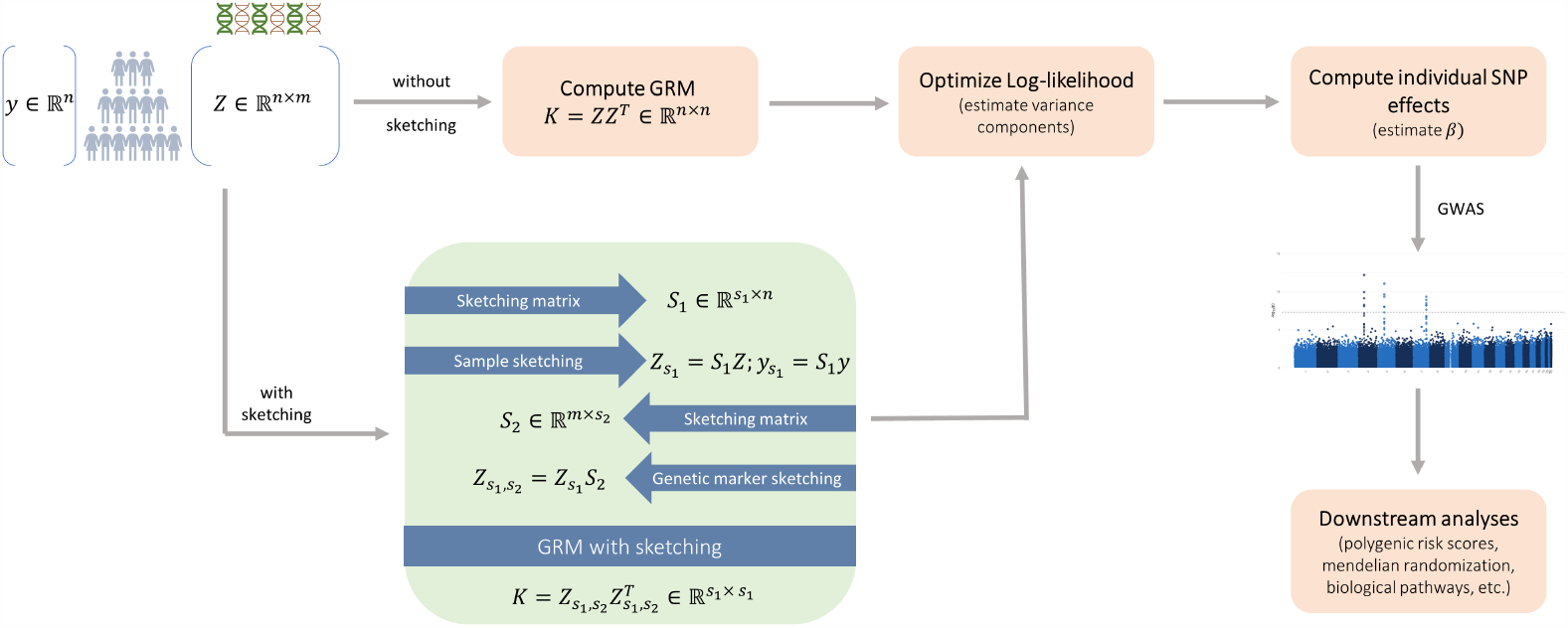
The MaSk-LMM framework. We use sketching to speed up the standard pipeline of LMM computations (peach). Our alternative pipeline uses sketching on both the sample and marker space of the genotype matrix **Z** (see eqns. (2) and (3)) to speed-up computations (green).

### 2.3 Data

Our experimental proof-of-principle evaluation seeks to demonstrate that sketching is a viable approach for LMMs. We chose to evaluate our algorithm on real and simulated data in order to show both run time and accuracy guarantees of MaSk-LMM when compared to current state-of-the-art.

#### Algorithm 1 MaSk-LMM

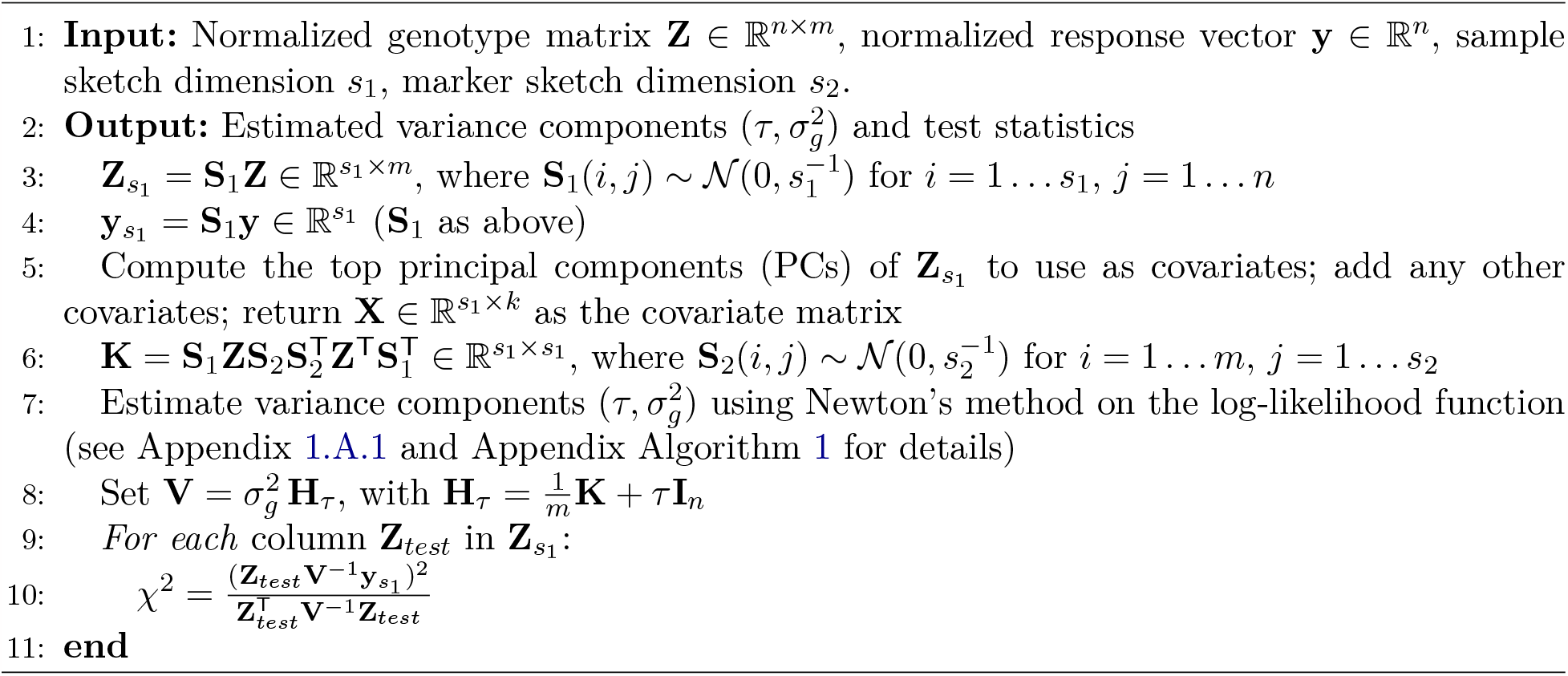

#### Simulated Genotypes

The synthetic data were generated from two ancestral backgrounds, Irish and British, using a “mosaic-chromosome” scheme modified from [18]. The general concept is to take a small set of individuals that are genetically distinct and generate artificial individuals by sampling their genomes. We began by selecting all individuals with British and Irish ancestries from the UK Biobank data after performing quality control and pruning, thus resulting in a dataset of 435,655 individuals and 265,642 SNPs. We then filtered the samples based on their ancestries inferred from SNP data (using the top two PCs, see Appendix Figure 1a) to ensure that the two groups were genetically distinct. We selected 100 samples from that subset of individuals to treat as the founders or ancestors to generate the artificial individuals from. We divided the genome into consecutive segments of 2,000 variants and generated *unrelated* individuals by selecting each segment from one of the 100 ancestors chosen at random and simulated *related* individuals by selecting the segments from a smaller number of ancestors according to the degree of relatedness. This process is done for both the Irish and British populations (see Appendix Figure 1b). Finally, we used GCTA tools [33] to simulate quantitative and binary traits for our simulated individuals.

#### Real Genotypes

The real genotypes were extracted from the UK Biobank for hypertension (HYP) and coronary artery disease (CAD). After performing quality control, the HYP dataset had 330k samples and 4.5M high quality SNPs (see Appendix 1.B.2 for details). The CAD dataset had 50k samples and 5.3M SNPs. The UKB datasets were created using a combination of NLP methods and manual curating to map ICD-10-CM codes to more meaningful phenotypes (see Appendix for details). We computed the top 20 principal components using TeraPCA [2].

## 3 Results

Our work focused on both theoretical and experimental properties of matrix sketching in the context of LMMs. From a theoretical perspective, we investigated the effect of marker sketching (using the matrix **S**_2_ of Section 2.2 and Algorithm 1) in downstream LMM computations. We leave the theoretical properties of using the sample sketching matrix **S**_1_ as an open problem for future research. From an experimental perspective, we evaluated the performance of MaSk-LMM on simulated and real-world genotypic datasets.

The experiments were performed at Purdue’s Negishi and Bell clusters, consisting of Dell compute nodes with two 64-core AMD Epyc 7662 Rome processors (128 cores per node) and 256 GB of memory. The nodes run CentOS 7 and use Slurm (Simple Linux Utility for Resource Management) as the batch scheduler for resource and job management.

### 3.1 Theoretical guarantees

A significant advantage of *matrix sketching* approaches is that they come with provable performance and accuracy guarantees. Indeed, this a major objective of our work: we provide a theoretical footing to our approach by proving that at least *marker sketching* (i.e., the use of the matrix **S**_2_ in eqns. (2) and (3)) results in bounded accuracy loss with high probability. The precise statement of our result appears in Theorem 3 in Appendix 1.B. Its proof uses a number of results from Randomized Linear Algebra along with information theoretic and probability theory inequalities.

We now present an informal statement of our results. In words, we prove that we can perform a binary hypothesis test on the parameters of an LMM as described in Section 2.1 and Appendix 1.A.1 by performing the computation on a marker-sketched version of the model. This sketching procedure only increases the error probability by a small constant *∈* that can be made arbitrarily small. Not surprisingly, the sketching dimension *s*_2_ depends on *∈*. Interestingly, the sketch dimension *s*_2_ depends linearly on *n* (the number of samples in the genotype matrix) and we also prove that this dependency is tight, i.e., it *can not be significantly reduced without catastrophically affecting the error*. We note again that this leaves as an open question the effect of sample sketching (namely, the use of the matrix **S**_1_ in eqns. (2) and (3)), which should be investigated in future work.

### 3.2 Experiments: Synthetic Data

For our experiments, we aimed to assess how MaSk-LMM performed in terms of execution times and accuracy of capturing causal associations (See Figure 2, Figure 3 and Table 2) when compared with other methods. These evaluations are key since *matrix sketching* at its core is an approximation and we need to practically evaluate its shortcomings. As shown in Table 2, we measured the average execution time of MaSk-LMM, BOLT-LMM, Regenie, and FaST-LMM when applied on our simulated datasets *D*_1_, *D*_2_, and *D*_3_. We used 10% as the sketch dimension for the samples (5% for *D*_3_) and 50% as the sketch dimension for the markers when calculating the GRM. As for the reasoning behind choosing these parameters, we selected them as to not be too aggressive using very small sketch dimensions (i.e. 1%) resulting in an inaccurate sketch, but also not using too high a sketch dimension (i.e. 80%) where we may just be introducing noise and not taking full advantage of the power of *matrix sketching*. We can see this tradeoff between accuracy and time in Figure 3. This choice may not be optimal for all datasets and should be tuned according to the number of samples and markers available. This is why we decided to use 5% sample sketching for *D*_3_ since we can still have enough samples for an accurate sketch (see Best Practices in Appendix 1.B.1 for more details and discussion).

**Table 1:** Real datasets (Coronary Artery Disease (CAD) and Hypertension (HYP)) and simulated datasets (*D*_1_, *D*_2_, *D*_3_).

| Dataset | Samples | SNPs | Size (.BED) |
| --- | --- | --- | --- |
| $D_1$ | 10,000 | 265,642 | 634 MB |
| $D_2$ | 100,000 | 265,642 | 6.2 GB |
| $D_3$ | 500,000 | 265,642 | 31 GB |
| CAD | 50,000 | 5,390,849 | 63 GB |
| HYP | 330,810 | 4,481,348 | 346 GB |

**Table 2:** Execution time (in minutes) of MaSk-LMM, Regenie, BOLT-LMM, and FaST-LMM when applied to the simulated datasets. Speed-up, in parentheses, achieved by MaSk-LMM compared to the other methods (* indicates no convergence after 50 hours, † indicates inability to allocate space for computation, ‡ indicates program-specific errors raised).

| Dataset | MaSk-LMM | Regenie | BOLT-LMM | FaST-LMM |
| --- | --- | --- | --- | --- |
| $D_1$ | 0.45 | 30.53 (67.84) | 22.00 (48.89) | 11.00 (24.44) |
| $D_2$ | 15.15 | 309.32 (20.42) | 219.63 (14.50) | n/a* (∞) |
| $D_3$ | 88.45 | 911.37 (10.30) | 1674.53 (18.93) | n/a*† (∞) |
| CAD | 34.1 | 188.6 (5.53) | n/a‡ (∞) | n/a*† (∞) |
| HYP | 268.2 | 956.1 (3.56) | n/a† (∞) | n/a*† (∞) |

**Fig. 2:**
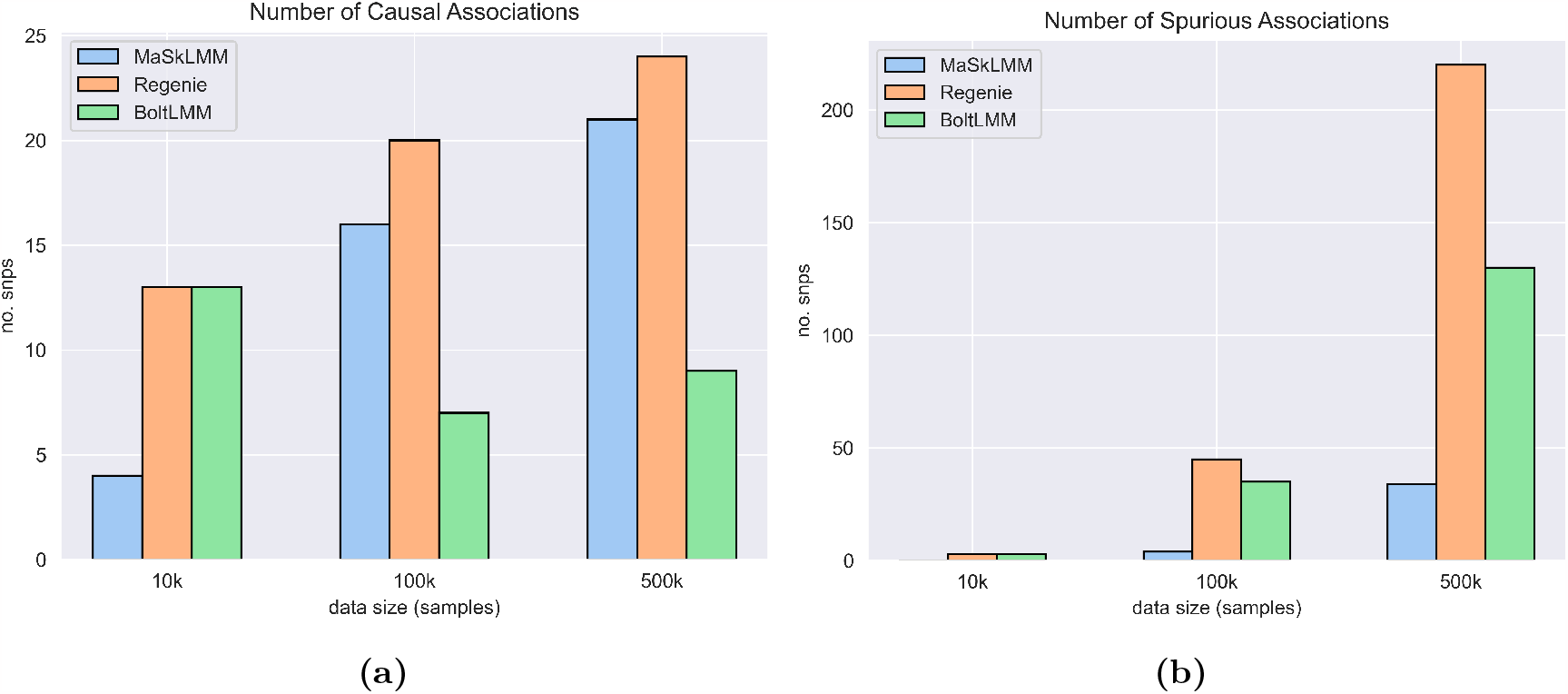
Average number of (a) causal and (b) spurious associations captured by MaSk-LMM, Regenie, and BOLT-LMM when applied to the British-Irish simulated data (265,462 SNPs and 10k, 100k, 500k samples). Software versions: Regenie, v3.2.5.3; BOLT-LMM v2.3

**Fig. 3:**
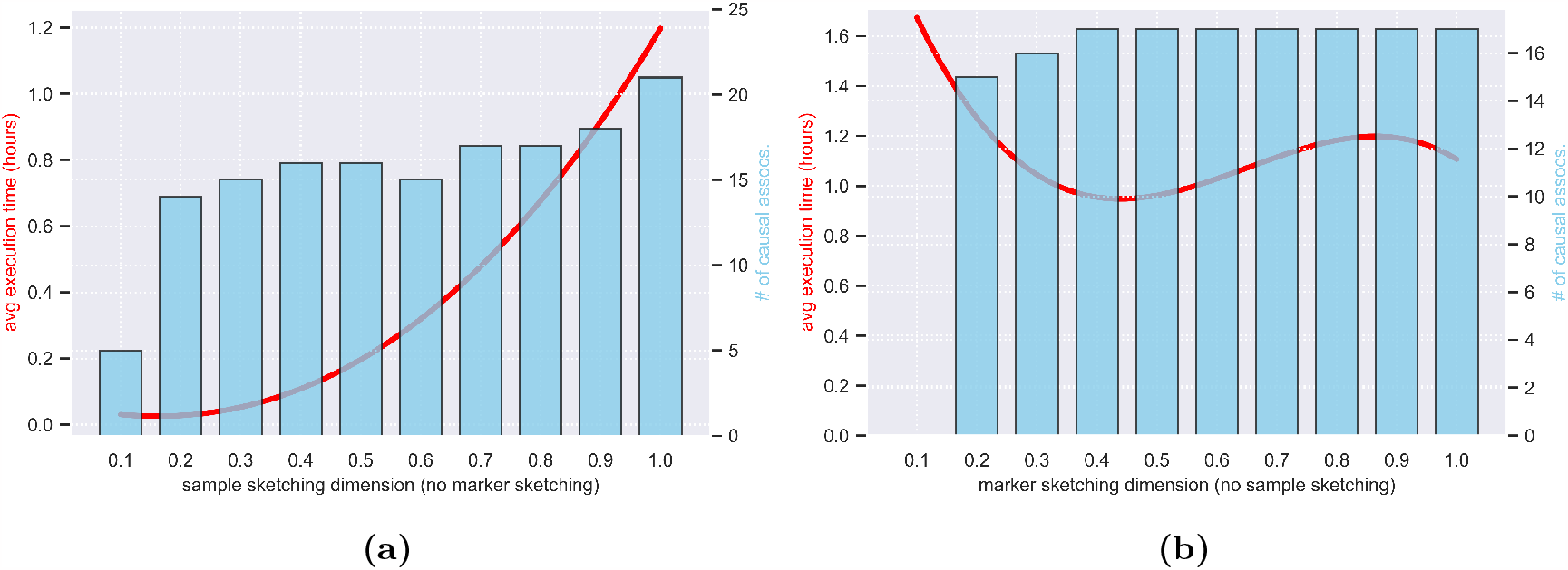
Number of causal associations and execution time of MaSk-LMM applied to *D*_1_ (British-Irish data with 10k samples and 265k SNPs) for varied sketch dimensions. (a) Applying no marker sketching and varying the sample sketching from 0.1 to 1.0. (b) Applying no sample sketching and varying the marker sketching from 0.1 to 1.0.

The results are the averages of 20 identical runs. MaSk-LMM achieved speed-ups in execution time of 49x, 15x and 19x over BOLT-LMM when run on *D*_1_, *D*_2_ and *D*_3_ respectively (Table 2). It also achieved speed-ups in execution time of 68x, 20x and 10x over Regenie when run on *D*_1_, *D*_2_ and *D*_3_ (Table 2). It also achieved a 24x speed-up over FaST-LMM when run on *D*_1_ (Table 2). FaST-LMM was unable to run on the other datasets in our computing environment. MaSk-LMM utilizes Newton’s method to estimate the parameters of the LMM and the number of iterations needed to converge can significantly impact the runtime and is largely dependent on the initial guess, which was set to 1.0. A better initial guess would result in even faster execution times and potentially more accurate solution. We also measured the average number of causal and spurious associations captured by MaSk-LMM, BOLT-LMM, and Regenie when applied on simulated datasets *D*_1_, *D*_2_, and *D*_3_ (see Figure 2 and Appendix Tables 1, 2, 3). Again, the results are the averages of 20 identical runs and we reported causal associations for each method using a *p*-value threshold of 1x10^−12^ to account for genome-wide significance. For each synthetic dataset, we simulated 25 markers as causal with a heritability ratio of 0.5 [33]. When applied to *D*_1_, MaSk-LMM performs worse than the other two methods despite being considerably faster. However, as we increase the sketch dimension, we do see improved performance with a tradeoff of longer running time (see Appendix Table 1). When applied to *D*_2_ and *D*_3_, MaSk-LMM outperforms BOLT-LMM, but is still slightly outperformed by Regenie in regards to capturing causal associations. However, MaSk-LMM captures fewer spurious associations in all scenarios compared to the other methods. We can see that our method steadily improves with respect to the number of causal associations that are captured as the data size grows, which illustrates the well-known fact that the performance and accuracy of matrix sketching improves when applied to larger datasets, especially when using smaller sketch dimensions [29].

### 3.3 Experiments: Real Data

We applied MaSk-LMM on datasets from complex disorders, including hypertension and coronary artery disease datasets. In both cases, MaSk-LMM identified biologically relevant associations efficiently.

#### Hypertension

We applied MaSk-LMM using a 5% sketch dimension for the samples and 50% sketch dimension for the markers on 330,810 individuals and 4,481,348 genotypes. We further improved the computational burden by generating the sketched input and GRM using the HYP dataset after pruning. MaSk-LMM identified 723 SNPs with a *p*-value threshold of 5x10^−8^ to account for genome-wide significance. We analyzed and assessed the significance of the associations by mapping them to diseases and disorders within the GWAS Catalog [24](Figure 4).

**Fig. 4:**
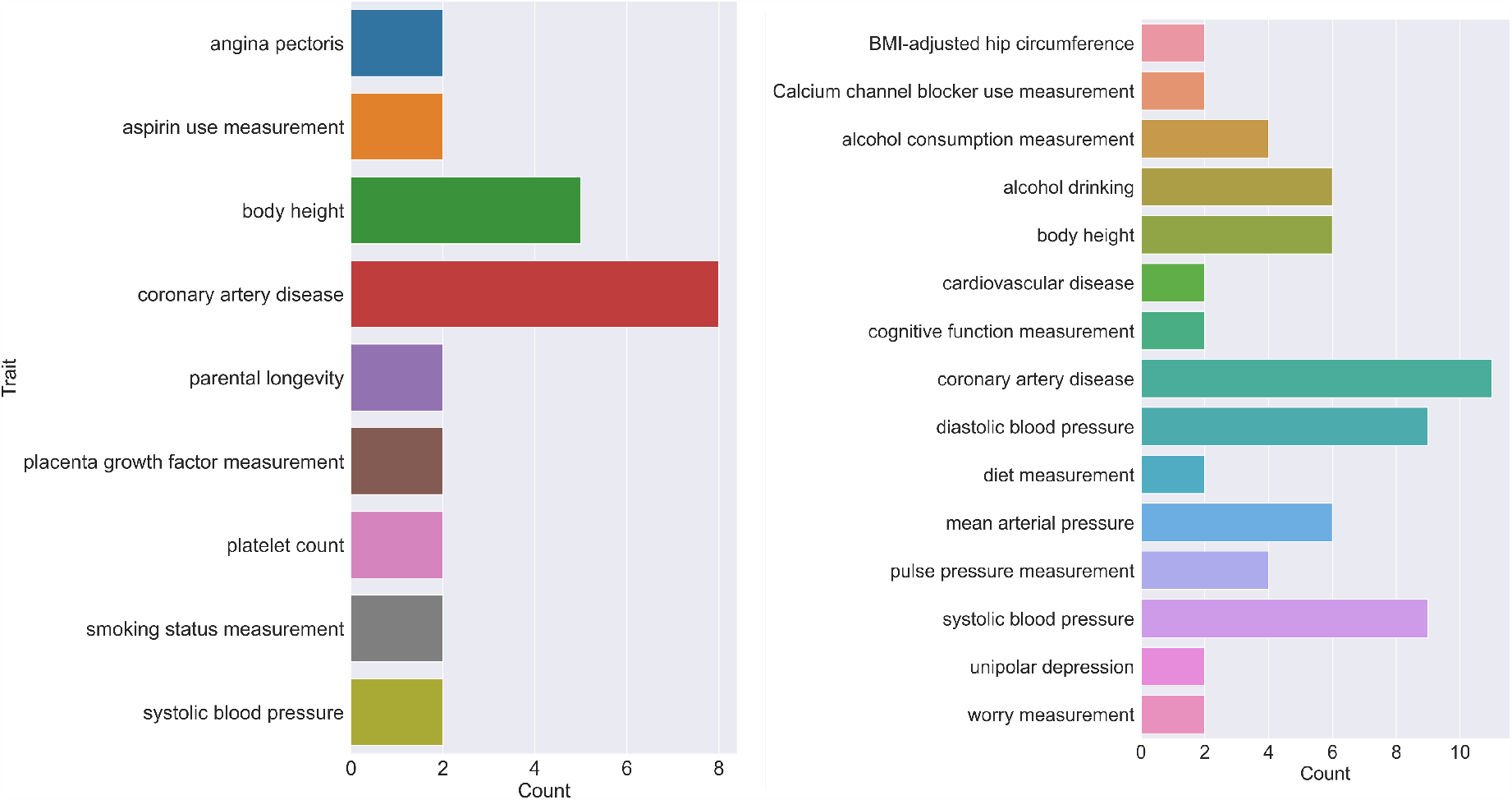
Bar chart of traits mapped to significant variants discovered by MaSkLMM for Coronary Artery Disease (left) and Hypertension (right) that have been validated in the GWAS Catalog [24].

We found some associations directly linked to *coronary artery disease* and many of the associations discovered by MaSk-LMM were connected to *systolic* and *diastolic blood pressure*. Elevated blood pressure represents a significant and controllable contributing factor to the development and progression of various clinical manifestations associated with coronary artery disease (CAD). The impact of high blood pressure extends across the spectrum of CAD-related conditions, making it a pivotal aspect in their pathogenesis. This condition underscores the importance of managing blood pressure as an integral part of preventing and managing coronary artery disease and its related health issues [28]. Additionally, thresholds between systolic and diastolic blood pressure are used to determine if a patient is hypertensive and their connection to cardiovascular outcomes continues to be studied [8]. Another significant finding was *mean arterial pressure* (MAP). MAP refers to the average of the arterial blood pressure in a single cardiac cycle. It can be an alternative index that can capture the overall exposure of an individual to increased pressure and be predictive of adverse events [14]. Other associations that MaSk-LMM discovered have well-established connections to hypertension such as *HDL cholesterol* and *alcohol consumption* [12, 27]. We attempted to compare the performance of Regenie and BOLT-LMM with MaSk-LMM when applied to the same dataset (see Table 2). BOLT-LMM was not able to allocate enough memory to run the program. In the case of Regenie, it discovered 4,493 SNPs above using the *p*-value threshold of 5x10^−8^. MaSk-LMM and Regenie had an overlap of 22 top associations. When increasing the sample sketch dimension to 10% and 20% for MaSk-LMM, the overlap increased to 340 and 655 top associations respectively. Regenie had a similar enrichment profile to MaSk-LMM finding strong connections to *systolic blood pressure, diastolic blood pressure*, and *mean arterial pressure*.

#### Coronary Artery Disease

We applied MaSk-LMM using a 10% sketch dimension for the samples and 50% sketch dimension for the markers on 50,000 individuals and 5,390,849 genotypes. We further improved the computational burden by generating the sketched input and GRM using the CAD dataset after pruning. MaSk-LMM identified 792 SNPs with a *p*-value threshold of 5x10^−8^ to account for genome-wide significance. We analyzed and assessed the significance of the associations by mapping them to diseases and disorders within the GWAS Catalog [24] (Figure 4). MaSk-LMM discovered many associations directly connected to *coronary artery disease*. Many other associations are strongly linked to adverse cardiovascular outcomes such as *angina pectoris, blood pressure*, and *myocardial infarction*. Angina pectoris arises when the myocardium (heart muscle) experiences insufficient blood and oxygen supply, a condition known as ischemia. It can manifest as a symptom of coronary artery disease (CAD). Studies continue to explore its connection to the clinical presentation and diagnosis of CAD [10]. Similar to hypertension, CAD and similar cardiovascular outcomes are heavily influenced by the relationship between systolic and diastolic blood pressure [8]. Lastly, myocardial infarction, commonly referred to as a “heart attack”, results from a reduction or complete halt in blood supply to a segment of the heart muscle, or myocardium. Myocardial infarctions can sometimes occur without noticeable symptoms, potentially going unnoticed, or they can manifest as a severe event causing a decline in heart function and unexpected fatality. The majority of myocardial infarctions are rooted in underlying coronary artery disease, which stands as the primary cause of mortality in the United States [4]. We attempted to compare the performance of Regenie and BOLT-LMM with MaSk-LMM when applied to the same dataset (see Table 2). BOLT-LMM was not able to allocate enough memory to run the program. In the case of Regenie, the program converged but did not capture any significant associations.

## 4 Discussion

We have developed a fast and efficient framework for linear mixed-model associations using matrix sketching. The resulting approach, MaSk-LMM, applies both sample and marker sketching to reduce the dimensions of the genotype matrix prior to performing LMM analysis. Such sketching speeds up the GRM computation as well as the estimation of the LMM parameters without a significant loss in accuracy. We presented theoretical results justifying the accuracy of sketching approaches in LMM computations. We also illustrated, using synthetic data, that our method runs faster than other state-of-the-art methods while capturing almost all of causal associations compared to the state-of-the-art methods (few, if any spurious associations are returned by MaSk-LMM). It is crucial to note that MaSk-LMM is a Python-based library whereas Regenie and BOLT-LMM are both written in C++. Studies have shown that C/C++ yields a better throughput with respect to memory usage and execution time [9]. For completeness, we compared MaSk-LMM with FaST-LMM [17], a Python-based tool implementing mixed models in association studies. MaSk-LMM significantly outperforms it in regards to execution time while still capturing significant associations (Table 2). We have further shown that MaSk-LMM can discover biologically relevant associations when applied to data for complex disorders like hypertension and coronary artery disease.

MaSk-LMM is an important advance and contribution to the space of genomics, specifically when conducting genome-wide association studies. Biobank-scale datasets spanning hundreds of thousands of individuals offer unprecedented opportunities to discover novel genetic loci associated with complex human traits and disease risk. However, they also present a computational challenge and burden. Using matrix sketching, we are able to harness the quality and richness that biobank-scale data offers while also alleviating the computational burden by reducing their dimensionality. While matrix sketching is a well-explored technique with robust theoretical underpinnings, its adoption in healthcare and life science applications remains limited. The primary reason for this limited acceptance is that the prevailing approach in these fields emphasizes accumulating ever-increasing volumes of data, while matrix sketching appears to diminish the data at first glance. However, we have demonstrated through the practical application of MaSk-LMM that matrix sketching can be a powerful and meaningful tool in this context. Our work with MaSk-LMM has showcased the potential and significance of matrix sketching in healthcare and life science applications. By embracing matrix sketching, we’ve managed to achieve significant benefits that mitigate the initial concerns about data reduction. This approach has opened new avenues for efficient data processing, analysis, and interpretation in these critical fields.

Even though MaSk-LMM is a powerful method in the space of LMMs and illustrates the power of approximate computations using matrix sketching, it is not without its limitations. First of all, there is a trade-off between the sketching dimension, the number of causal associations captured, and its running time (see Appendix 1.B.1 and Appendix Tables 4 and 5). Using more aggressive sketching and reducing the number of retained markers or samples (parameters *s*_1_ and *s*_2_ in Algorithm 1) to 5-10% of the original values *m* and *n*, reduces the running time but significantly worsens the quality of the approximation, resulting in fewer causal associations captured and potentially more spurious associations. This issue becomes less prevalent as the dataset size increases, since the abundance of markers and samples help improve the quality of the sketching when using smaller sketch dimensions. Additionally, our current implementation has not incorporated the leave-one-chromosome-out cross-validation (LOCO) to correct for proximal contamination, a phenomenon that might result in loss of power if the candidate marker is included in the GRM [34]. However, in our setting, the input is sketched and the GRM computation operates on a much smaller matrix, which seems to mitigate this issue, at least in our empirical evaluations. Other future research directions that could improve our framework include taking advantage of sparsity in our computations, improving data management, as well as implementing our methods in an environment that is more suitable for high-performance with biobank-scale data, like C++ with Intel’s OpenMPI supporting libraries.

## Data Availability

Data analysis was performed under UK Biobank application 50658 using existing publicly available and deidentified data and was IRB exempt.

## Code Availability

A Python implementation of MaSK-LMM is available at: https://github.com/IBM/mask-lmm.

## Footnotes

3 We use bold letters for vectors and matrices; a vector **x** ∈ ℝ^*n*^ is an *n*-dimensional real vector, while a matrix **X** *œ* ℝ ∈^*n×m*^ is an *n × m* real matrix.

4 We use the notation *𝒩* (*μ*, Σ) to denote a multivariate normal distribution with mean vector *μ* and covariance matrix Σ. **I**_*n*_ denotes the *n × n* identity matrix.

