## Supplementary Material for "MaSk-LMM: A Matrix Sketching Framework for Linear Mixed Models in Association Studies"

### Appendix 1.A

#### 1.A.1 LMM Foundations

In this section, we use $\mathbf{I}_n$ to denote the $n \times n$ identity matrix; $\mathbf{0}$ to denote the all-zeros matrix or vector, with appropriate subscripts (if needed) to indicate dimensions; $\det(\mathbf{X})$ denotes the determinant of the matrix $\mathbf{X}$; and $\mathrm{tr}(\mathbf{X})$ denotes the trace of the matrix $\mathbf{X}$. Recall that a two component genomic variance model for linear mixed models is typically described in the following form:

$$\mathbf{y} = \mathbf{X}\boldsymbol{\beta} + \frac{1}{\sqrt{m}}\mathbf{Z}\mathbf{u} + \mathbf{e},$$

where $\mathbf{y} \in \mathbb{R}^n$ is the measured phenotype (response); $\mathbf{X} \in \mathbb{R}^{n\times k}$ is the matrix of the $k$ covariates (*e.g.* principal components, age, sex, etc.) with the corresponding vector of fixed effects $\boldsymbol{\beta} \in \mathbb{R}^n$; $\mathbf{Z} \in \mathbb{R}^{n\times m}$ is the genotype matrix of $n$ individuals genotyped on $m$ genetic markers with $\mathbf{u} \in \mathbb{R}^m$ being the corresponding genetic effects vector; and $\mathbf{e} \in \mathbb{R}^n$ is the error vector or the component of $\mathbf{y}$ which cannot be explained by the model. We assume $\mathbf{u}$ and $\mathbf{e}$ are independent vectors and moreover that $\mathbf{u} \sim \mathcal{N}\left(\mathbf{0}, \sigma_g^2\,\mathbf{I}_m\right)$ and $\mathbf{e} \sim \mathcal{N}\left(\mathbf{0}, \sigma_e^2\,\mathbf{I}_n\right)$ with scalars $\sigma_g^2$ and $\sigma_e^2$ being the heritable and non-heritable components of $\mathbf{u}$ and $\mathbf{e}$ respectively. In the LMM setting, some form of maximum likelihood estimation is used to estimate the random and fixed effects components of the model in order to identify genetic associations while correcting for confounding effects. Define

$$\mathbf{H}_\tau = \frac{1}{m}\mathbf{Z}\mathbf{Z}^\top + \tau\mathbf{I}_n. \tag{1}$$

Note that as $\mathbf{y}$ is a linear transformation[3] of the independent multivariate normal random vectors $\mathbf{u}$ and $\mathbf{e}$, it is also multivariate normal and in fact,

$$\mathbf{y} \sim \mathcal{N}(\mathbf{X}\boldsymbol{\beta}, \sigma_g^2\,\mathbf{H}_\tau)\,,$$

where $\tau = \frac{\sigma_e^2}{\sigma_g^2}$. Let $\mathbf{U}_{\mathbf{X},\perp} \in \mathbb{R}^{n\times(n-k)}$ be a projection to a $(n-k)$-dimensional subspace that is perpendicular to the columns of $\mathbf{X}$ *i.e.* $\mathbf{U}_{\mathbf{X},\perp}^\top\mathbf{X} = \mathbf{0}$. So, we can rewrite the model by pre-multiplying both sides by $\mathbf{U}_{\mathbf{X},\perp}^\top$ to get

$$\mathbf{U}_{\mathbf{X},\perp}^\top\mathbf{y} = \frac{1}{\sqrt{m}}\mathbf{U}_{\mathbf{X},\perp}^\top\mathbf{Z}\mathbf{u} + \mathbf{U}_{\mathbf{X},\perp}^\top\mathbf{e}\,. \tag{2}$$

[3] Recall that if $\mathbf{p} \sim \mathcal{N}(\mu_\mathbf{p}, \Sigma_\mathbf{p})$, then $\mathbf{A}\mathbf{p} + \mathbf{q} \sim \mathcal{N}(\mathbf{A}\mu_\mathbf{p}, \mathbf{A}\Sigma_\mathbf{p}\mathbf{A}^\top)\,.$

Clearly, $\mathbf{U}_{\mathbf{X},\perp}^{\mathsf{T}}\mathbf{y}$ is also a linear transformation on $\mathbf{y}$ and therefore, we have $\mathbf{U}_{\mathbf{X},\perp}^{\mathsf{T}}\mathbf{y} \sim \mathcal{N}(\mathbf{0}, \sigma_g^2\, \mathbf{U}_{\mathbf{X},\perp}^{\mathsf{T}}\mathbf{H}_\tau\mathbf{U}_{\mathbf{X},\perp})$. Now, given the data, we express the likelihood function as follows

$$L(\tau, \sigma_g^2 \mid \mathbf{U}_{\mathbf{X},\perp}^{\mathsf{T}}\mathbf{y}) = \frac{\exp\left(-\frac{1}{2\sigma_g^2}\,\mathbf{y}^{\mathsf{T}}\mathbf{U}_{\mathbf{X},\perp}(\mathbf{U}_{\mathbf{X},\perp}^{\mathsf{T}}\mathbf{H}_\tau\mathbf{U}_{\mathbf{X},\perp})^{-1}\mathbf{U}_{\mathbf{X},\perp}^{\mathsf{T}}\mathbf{y}\right)}{(2\pi)^{\frac{n-k}{2}}\cdot\left(\det(\sigma_g^2\,\mathbf{U}_{\mathbf{X},\perp}^{\mathsf{T}}\mathbf{H}_\tau\mathbf{U}_{\mathbf{X},\perp})\right)^{1/2}}. \tag{3}$$

We now get the log-likelihood just by taking the log on the both sides of eqn. (3) as follows:

$$\begin{aligned}\ell(\tau, \sigma_g^2 \mid \mathbf{U}_{\mathbf{X},\perp}^{\mathsf{T}}\mathbf{y}) = -\frac{n-k}{2}\log 2\pi - \frac{n-k}{2}\log\sigma_g^2 - \frac{1}{2}\log\left(\det(\mathbf{U}_{\mathbf{X},\perp}^{\mathsf{T}}\mathbf{H}_\tau\mathbf{U}_{\mathbf{X},\perp})\right)\\ - \frac{1}{2\sigma_g^2}\cdot\mathbf{y}^{\mathsf{T}}\mathbf{U}_{\mathbf{X},\perp}(\mathbf{U}_{\mathbf{X},\perp}^{\mathsf{T}}\mathbf{H}_\tau\mathbf{U}_{\mathbf{X},\perp})^{-1}\mathbf{U}_{\mathbf{X},\perp}^{\mathsf{T}}\mathbf{y}.\end{aligned} \tag{4}$$

Differentiation of eqn. (4) w.r.t $\sigma_g^2$ and $\tau$ yields

$$\frac{\partial\ell}{\partial\sigma_g^2} = -\frac{n-k}{2\sigma_g^2} + \frac{1}{2\sigma_g^4}\,\mathbf{y}^{\mathsf{T}}\mathbf{U}_{\mathbf{X},\perp}(\mathbf{U}_{\mathbf{X},\perp}^{\mathsf{T}}\mathbf{H}_\tau\mathbf{U}_{\mathbf{X},\perp})^{-1}\mathbf{U}_{\mathbf{X},\perp}^{\mathsf{T}}\mathbf{y} \tag{5a}$$

$$\frac{\partial\ell}{\partial\tau} = -\frac{1}{2}\cdot\frac{\partial\mathit{\Delta}_1}{\partial\tau} - \frac{1}{2\sigma_g^2}\cdot\frac{\partial\mathit{\Delta}_2}{\partial\tau}, \tag{5b}$$

where $\mathit{\Delta}_1 = \log\left(\det(\mathbf{U}_{\mathbf{X},\perp}^{\mathsf{T}}\mathbf{H}_\tau\mathbf{U}_{\mathbf{X},\perp})\right)$ and $\mathit{\Delta}_2 = \mathbf{y}^{\mathsf{T}}\mathbf{U}_{\mathbf{X},\perp}(\mathbf{U}_{\mathbf{X},\perp}^{\mathsf{T}}\mathbf{H}_\tau\mathbf{U}_{\mathbf{X},\perp})^{-1}\mathbf{U}_{\mathbf{X},\perp}^{\mathsf{T}}\mathbf{y}$. Next, we differentiate each of the terms on the right hand side of eqn. (5b) separately. First,

$$\begin{aligned}\frac{\partial\mathit{\Delta}_1}{\partial\tau} &= \frac{\partial\log\left(\det(\mathbf{U}_{\mathbf{X},\perp}^{\mathsf{T}}\mathbf{H}_\tau\mathbf{U}_{\mathbf{X},\perp})\right)}{\partial\det(\mathbf{U}_{\mathbf{X},\perp}^{\mathsf{T}}\mathbf{H}_\tau\mathbf{U}_{\mathbf{X},\perp})}\cdot\frac{\partial\det(\mathbf{U}_{\mathbf{X},\perp}^{\mathsf{T}}\mathbf{H}_\tau\mathbf{U}_{\mathbf{X},\perp})}{\partial\tau}\\ &= \frac{1}{\det(\mathbf{U}_{\mathbf{X},\perp}^{\mathsf{T}}\mathbf{H}_\tau\mathbf{U}_{\mathbf{X},\perp})}\cdot\det(\mathbf{U}_{\mathbf{X},\perp}^{\mathsf{T}}\mathbf{H}_\tau\mathbf{U}_{\mathbf{X},\perp})\ \mathrm{tr}\left((\mathbf{U}_{\mathbf{X},\perp}^{\mathsf{T}}\mathbf{H}_\tau\mathbf{U}_{\mathbf{X},\perp})^{-1}\frac{\partial(\mathbf{U}_{\mathbf{X},\perp}^{\mathsf{T}}\mathbf{H}_\tau\mathbf{U}_{\mathbf{X},\perp})}{\partial\tau}\right)\\ &= \mathrm{tr}\left((\mathbf{U}_{\mathbf{X},\perp}^{\mathsf{T}}\mathbf{H}_\tau\mathbf{U}_{\mathbf{X},\perp})^{-1}\frac{\partial\left(\frac{1}{m}\mathbf{U}_{\mathbf{X},\perp}^{\mathsf{T}}\mathbf{Z}\mathbf{Z}^{\mathsf{T}}\mathbf{U}_{\mathbf{X},\perp} + \tau\mathbf{I}_{n-k}\right)}{\partial\tau}\right) = \mathrm{tr}\left((\mathbf{U}_{\mathbf{X},\perp}^{\mathsf{T}}\mathbf{H}_\tau\mathbf{U}_{\mathbf{X},\perp})^{-1}\right),\end{aligned} \tag{6}$$

where the second equality follows from $\frac{\partial\,\det(\mathbf{A})}{\partial x} = \det(\mathbf{A})\,\mathrm{tr}\left(\mathbf{A}^{-1}\frac{\partial\mathbf{A}}{\partial x}\right)$. Similarly for the second term in eqn. (5b), we have

$$\begin{aligned}\frac{\partial\mathit{\Delta}_2}{\partial\tau} &= \frac{\partial\left(\mathbf{y}^{\mathsf{T}}\mathbf{U}_{\mathbf{X},\perp}(\mathbf{U}_{\mathbf{X},\perp}^{\mathsf{T}}\mathbf{H}_\tau\mathbf{U}_{\mathbf{X},\perp})^{-1}\mathbf{U}_{\mathbf{X},\perp}^{\mathsf{T}}\mathbf{y}\right)}{\partial\tau} = \mathbf{y}^{\mathsf{T}}\mathbf{U}_{\mathbf{X},\perp}\left(\frac{\partial(\mathbf{U}_{\mathbf{X},\perp}^{\mathsf{T}}\mathbf{H}_\tau\mathbf{U}_{\mathbf{X},\perp})^{-1}}{\partial\tau}\right)\mathbf{U}_{\mathbf{X},\perp}^{\mathsf{T}}\mathbf{y}\\ &= \mathbf{y}^{\mathsf{T}}\mathbf{U}_{\mathbf{X},\perp}\left(-(\mathbf{U}_{\mathbf{X},\perp}^{\mathsf{T}}\mathbf{H}_\tau\mathbf{U}_{\mathbf{X},\perp})^{-1}\left(\frac{\partial(\mathbf{U}_{\mathbf{X},\perp}^{\mathsf{T}}\mathbf{H}_\tau\mathbf{U}_{\mathbf{X},\perp})}{\partial\tau}\right)(\mathbf{U}_{\mathbf{X},\perp}^{\mathsf{T}}\mathbf{H}_\tau\mathbf{U}_{\mathbf{X},\perp})^{-1}\right)\mathbf{U}_{\mathbf{X},\perp}^{\mathsf{T}}\mathbf{y}\\ &= -\,\mathbf{y}^{\mathsf{T}}\mathbf{U}_{\mathbf{X},\perp}(\mathbf{U}_{\mathbf{X},\perp}^{\mathsf{T}}\mathbf{H}_\tau\mathbf{U}_{\mathbf{X},\perp})^{-2}\mathbf{U}_{\mathbf{X},\perp}^{\mathsf{T}}\mathbf{y} = -\left\|(\mathbf{U}_{\mathbf{X},\perp}^{\mathsf{T}}\mathbf{H}_\tau\mathbf{U}_{\mathbf{X},\perp})^{-1}\mathbf{U}_{\mathbf{X},\perp}^{\mathsf{T}}\mathbf{y}\right\|_2^2,\end{aligned} \tag{7}$$

where the second equality holds as $\frac{\partial\mathbf{A}^{-1}}{\partial x} = -\mathbf{A}^{-1}\frac{\partial\mathbf{A}}{\partial x}\mathbf{A}^{-1}$ and the last equality directly follows from eqn. (6) that $\frac{\partial\left(\mathbf{U}_{\mathbf{X},\perp}^{\top}\mathbf{H}_\tau\mathbf{U}_{\mathbf{X},\perp}\right)}{\partial\tau} = \mathbf{I}_{n-k}$. Finally, combining eqns. (6) and (7), we rewrite eqn. (5b) as follows

$$\frac{\partial\ell}{\partial\tau} = -\frac{1}{2}\ \mathrm{tr}\left(\left(\mathbf{U}_{\mathbf{X},\perp}^{\top}\mathbf{H}_\tau\mathbf{U}_{\mathbf{X},\perp}\right)^{-1}\right) + \frac{1}{2\sigma_g^2}\left\|\left(\mathbf{U}_{\mathbf{X},\perp}^{\mathsf{T}}\mathbf{H}_\tau\mathbf{U}_{\mathbf{X},\perp}\right)^{-1}\mathbf{U}_{\mathbf{X},\perp}^{\top}\mathbf{y}\right\|_2^2. \tag{8}$$

Equating eqns. (5a) and (8) to zero gives the REML estimators. The resulting equations clearly have no analytic solution and have to be solved numerically. A standard iterative procedure [2,4,6] (*i.e.* Newton's method) is to first assign initial values to $\tau$ and then (i) solve

$$\hat{\sigma}_g^2 = \frac{\mathbf{y}^\top \mathbf{U}_{\mathbf{X},\perp} \left(\mathbf{U}_{\mathbf{X},\perp}^\top \mathbf{H}_\tau \mathbf{U}_{\mathbf{X},\perp}\right)^{-1} \mathbf{U}_{\mathbf{X},\perp}^\top \mathbf{y}}{n-k} \tag{9}$$

based on eqn. (5a), and (ii) use the $\tau$ and $\hat{\sigma}_g^2$ from eqn. (9) to calculate new $\tau$ that makes eqn. (8) closer to zero. Repetition of (i) and (ii), ending at (i), is continued until a desired degree of accuracy is attained.

**Computing $\mathbf{U}_{\mathbf{X},\perp}$.** One way to compute $\mathbf{U}_{\mathbf{X},\perp}$ to use a QR decomposition on the matrix of covariates $\mathbf{X}$. Then, the matrix $\mathbf{Q} \in \mathbb{R}^{n\times k}$ would be a basis for the column span of $\mathbf{X}$. Then, it follows that

$$\mathbf{U}_{\mathbf{X},\perp}\mathbf{U}_{\mathbf{X},\perp}^T = \mathbf{I} - \mathbf{Q}\mathbf{Q}^T = \mathbf{I} - \mathbf{U}_\mathbf{X}\mathbf{U}_\mathbf{X}^T.$$

**Newton's Method.** As stated above, the system of equations emerging from the log-likelihood estimation have no analytic solution and need to be solved numerically. We chose to use a standard iterative approach (Newton's method) to estimate the parameters. More specifically, we use the secant method which is a finite-difference approximation of Newton's method.

**Algorithm 1** Newton's Method to estimate variance components of LMM

**Input:** Sketched response vector $\mathbf{y}_{s_1} \in \mathbb{R}^n$, sketched GRM matrix $\mathbf{K} \in \mathbb{R}^{s_1\times s_1}$, number of samples $n$, number of markers $m$, number of covariates $k$, tolerance for iterative method $tol$, initial guess $\tau_0$.
**Output:** Estimated variance components $(\tau_0, \sigma_g^2)$
$newton\_raphson(\mathbf{y}_{s_1}, \mathbf{K}, \mathbf{U}_\mathbf{X}, n, m, k, tol, \tau_0)$ :
  $\mathbf{H}_\tau = \frac{1}{m}\mathbf{K} + \tau_0 \mathbf{I}_n$
  $\mathbf{P} = (\mathbf{I} - \mathbf{U}_\mathbf{X}\mathbf{U}_\mathbf{X}^\top)\mathbf{H}_\tau(\mathbf{I} - \mathbf{U}_\mathbf{X}\mathbf{U}_\mathbf{X}^\top)$
  $\mathbf{P}_{inv} = \mathbf{P}^\dagger$
  $\sigma_g^2 = \frac{1}{n-k}\mathbf{y}_{s_1}^\top \mathbf{P}_{inv} \mathbf{y}_{s_1}$
  $lle = -\frac{n-k}{2}\log 2\pi - \frac{n-k}{2}\log \sigma_g^2 - \frac{1}{2}\log(\det(\mathbf{P})) - \frac{n-k}{2}$
  Use secant method on $lle$ to determine current $\delta$ (convergence criterion)
  if $|\delta| < tol$ :
    **return** $\tau_0, \sigma_g^2$
  else:
    **return** $newton\_raphson(\mathbf{y}_{s_1}, \mathbf{K}, \mathbf{U}_\mathbf{X}, n, m, k, tol, \tau_0 - \delta)$
**end**

### Appendix 1.B Our theoretical contributions

In this section, we show that sketching the random effects matrix $\mathbf{Z} \in \mathbb{R}^{n\times m}$ of an LMM described in Section 2.1 maintains enough information for effective statistical inference. Specifically, we focus on the binary testing problem for an LMM, where we decide between two parameter sets, $(\sigma_{g,0}^2, \tau_0)$ and $(\sigma_{g,1}^2, \tau_1)$. We show that by replacing the random effects matrix in an LMM[4] $\mathbf{ZS}$, where $\mathbf{S} \in \mathbb{R}^{m\times s}$ is

[4] For notational simplicity, in this section we drop the subscript in the marker sketching matrix $\mathbf{S}_2$ and denote it by just $\mathbf{S}$. We also drop the subscript in the marker sketching dimension $s_2$ and denote it by just $s$.

a Gaussian sketching matrix and $s$ is linear in $n$, the original problem can be decided with arbitrarily small increase in the testing error by instead deciding the sketched problem. See Theorem 3 for a precise statement of our results.

Let $\mathbb{P}_{\mathcal{D}}(\cdot)$ denote the probability of an event under the distribution $\mathcal{D}$. We will use results about the following random matrix distribution.

**Definition 1.** ***(Wishart Ensemble)*** *An $n \times n$ matrix $\mathbf{W}$ is sampled from a Wishart ensemble with $p$ degrees of freedom and covariance matrix $\mathbf{\Sigma}$ (denoted $\mathbf{W} \sim \mathbf{W}_n(p, \mathbf{\Sigma})$) if $\mathbf{W} = \mathbf{G}\mathbf{G}^T$, where $\mathbf{G} \in \mathbb{R}^{n \times p}$ and each row of $\mathbf{G}$ is distributed i.i.d. as $\mathbf{G}_{i*} \sim \mathcal{N}(\mathbf{0}_p, \mathbf{\Sigma})$.*

Next, we introduce some necessary concepts from information theory. For more information on the below definitions and theorems, we refer the reader to [7].

**Definition 2.** ***(KL-Divergence)*** *Given probability distributions $\mathcal{P}$ and $\mathcal{Q}$ both supported on the set $\mathcal{X}$ with probability density functions $p(\cdot)$ and $q(\cdot)$ respectively, the KL-divergence from $\mathcal{Q}$ to $\mathcal{P}$ is denoted by:*

$$\mathrm{D}_{\mathrm{KL}}(\mathcal{P}||\mathcal{Q}) = \int_{\mathcal{X}} p(x) \log_2 \frac{p(x)}{q(x)}.$$

*If there exists $x \in \mathcal{X}$ such that $q(x) = 0$ and $p(x) \neq 0$ or vice-versa, then the KL-divergence is defined to be infinite.*

**Definition 3.** ***(Total Variation Distance)*** *Given probability distributions $\mathcal{P}$ and $\mathcal{Q}$ both supported on the set $\mathcal{X}$, the total variation distance is denoted by:*

$$D_{TV}(\mathcal{P}, \mathcal{Q}) = \sup_{X \subset \mathcal{X}} \left| \mathbb{P}_{\mathcal{P}}(X) - \mathbb{P}_{\mathcal{Q}}(X) \right|.$$

The so-called "data processing inequality" is stated in many forms, but the following will be most useful for our purposes. It will allow us to bound the KL-divergence between two distributions by bounding the KL-divergence between a different pair of distributions.

**Theorem 1.** ***(Data processing inequality)*** *Let $\mathcal{P}$ and $\mathcal{Q}$ be probability distributions with probability density functions, and let $f(\cdot)$ be an arbitrary (possibly random) function where the randomness of $f(\cdot)$ is independent from $\mathcal{P}$ and $\mathcal{Q}$. Then, the following inequality holds,*

$$\mathrm{D}_{\mathrm{KL}}(f(\mathcal{P}) \ || \ f(\mathcal{Q})) \leq \mathrm{D}_{\mathrm{KL}}(\mathcal{P} \ || \ \mathcal{Q}),$$

*where $f(\mathcal{P})$ and $f(\mathcal{Q})$ denotes the distribution of $f(\cdot)$ applied to the random variables with the respective distributions.*

The following well-known inequality allows us bound total variation distance between two distributions by the KL-divergence.

**Theorem 2.** ***(Pinsker's inequality)*** *Let $\mathcal{P}$ and $\mathcal{Q}$ be probability distributions that have corresponding probability density functions. Then,*

$$D_{TV}(\mathcal{P}, \mathcal{Q}) \leq \sqrt{\frac{1}{2} \mathrm{D}_{\mathrm{KL}}(\mathcal{P}||\mathcal{Q})}.$$

First, we start by bounding the KL-divergence between zero-centered multivariate Gaussian distributions with differing covariance matrices. The bound in KL-divergence between these distributions will later be used to bound the KL-divergence between different LMMs via the data-processing inequality.

**Lemma 1.** *Let* $\mathcal{D}_1 = \mathcal{N}(\mathbf{0}, \mathbf{Z}\mathbf{Z}^T)$ *and* $\mathcal{D}_2 = \mathcal{N}(\mathbf{0}, \mathbf{Z}\mathbf{S}\mathbf{S}^T\mathbf{Z}^T)$*, where* $\mathbf{Z} \in \mathbb{R}^{n\times m}$ *is a fixed full-rank matrix with* $m > n$ *and* $\mathbf{S} \in \mathbb{R}^{m\times s}$ *is a Gaussian sketching matrix. i.e., each entry of* $\mathbf{S}$ *is i.i.d. as* $\mathbf{S}_{ij} \sim \mathcal{N}(0, 1/s)$*. If* $s = \mathcal{O}\left(\frac{n}{\epsilon^2}\right)$*, then for large enough* $n$*, with probability at least 0.98,*

$$\mathrm{D}_{\mathrm{KL}}(\mathcal{D}_1||\mathcal{D}_2) \leq \epsilon.$$

*Proof.* By Section 9 of [3], we have the following closed form expression for the KL-divergence between $\mathcal{D}_1$ and $\mathcal{D}_2$:

$$\mathrm{D}_{\mathrm{KL}}(\mathcal{D}_2||\mathcal{D}_1) = \frac{1}{2}\left(\log\frac{\det \mathbf{Z}\mathbf{Z}^T}{\det \mathbf{Z}\mathbf{S}\mathbf{S}^T\mathbf{Z}^T} - n + \mathrm{tr}[(\mathbf{Z}\mathbf{Z}^T)^{-1}(\mathbf{Z}\mathbf{S}\mathbf{S}^T\mathbf{Z}^T)]\right).$$

We will bound the KL-divergence between $\mathcal{D}_1$ and $\mathcal{D}_2$ with high probability over the distribution of the sketching matrix $\mathbf{S}$. We first bound the log-determinant term.

Note that each row of $\mathbf{S}^T\mathbf{Z}^T$ is independently distributed as $[\mathbf{S}^T\mathbf{Z}^T]_i \sim \mathcal{N}(\mathbf{0}, \mathbf{Z}\mathbf{Z}^T)$. Hence, $\mathbf{Z}\mathbf{S}\mathbf{S}^T\mathbf{Z}^T$ is distributed as a Wishart ensemble. By Theorem 1 in [1], if $\frac{n}{s} \to \eta \in (0, 1)$,

$$\frac{\log\det \mathbf{Z}\mathbf{S}\mathbf{S}^T\mathbf{Z}^T - \sum_{i=1}^n \log\left(1 - \frac{i}{s-1}\right) - \log\det \mathbf{Z}\mathbf{Z}^T}{\sqrt{-2\log\left(1 - \frac{n}{s}\right)}} \to^L \mathcal{N}(0, 1) \text{ as } s \to \infty,$$

where $\to^L$ denote convergence in distribution. For fixed $\eta$, we can simplify the summation term as follows:

$$\sum_{i=1}^n \log\left(1 - \frac{i}{s-1}\right) \to \int_0^\eta \log(1-x)\,dx = (\eta - 1)\log(1-\eta) - \eta, \qquad \text{as } n \to \infty,\ \frac{n}{s} \to \eta.$$

Therefore, for large $n$ and $s$, we have the following asymptotic equality in distribution (denoted by $\approx_D$), where $g \sim \mathcal{N}(0, 1)$,

$$\left|\log\det \mathbf{Z}\mathbf{S}\mathbf{S}^T\mathbf{Z}^T - \log\det \mathbf{Z}\mathbf{Z}^T\right| \approx_D \sqrt{-2\log(1-\eta)} \cdot g + (\eta - 1)\log(1-\eta) - \eta. \tag{10}$$

Using the inequality $\log(x) \geq 2x - 2$ for $x \in [1/2, 1]$, we can solve for a value of $\eta \in (0, 1/2)$ that guarantees that $|\log\det \mathbf{Z}\mathbf{S}\mathbf{S}^T\mathbf{Z}^T - \log\det \mathbf{Z}\mathbf{Z}^T|$ is bounded by some $\epsilon \in (0, 1)$. With probability 0.99, $|g| \leq 3$. Hence, with probability at least 0.99,

$$\begin{aligned}
|\log\det \mathbf{Z}\mathbf{S}\mathbf{S}^T\mathbf{Z}^T - \log\det \mathbf{Z}\mathbf{Z}^T| &\approx_D \sqrt{-2\log(1-\eta)} \cdot g + (\eta - 1)\log(1-\eta) - \eta \\
&\leq \sqrt{4\eta} \cdot 3 + 2(1-\eta)\eta - \eta \\
&\leq C\sqrt{\eta},
\end{aligned}$$

where $C$ is some universal constant. Therefore, $\eta = \mathcal{O}(\epsilon^2)$ suffices to guarantee that

$$|\log\det \mathbf{Z}\mathbf{S}\mathbf{S}^T\mathbf{Z}^T - \log\det \mathbf{Z}\mathbf{Z}^T| < \epsilon$$

in the previous inequality asymptotically with probability at least 0.99. While eqn. (10) only denotes asymptotic equality in distribution, convergence in distribution implies that the probability that

$$|\log\det \mathbf{Z}\mathbf{S}\mathbf{S}^T\mathbf{Z}^T - \log\det \mathbf{Z}\mathbf{Z}^T| > C\sqrt{\eta}$$

holds differs from the asymptotic result by an arbitrarily small amount for large enough $n$. Therefore, for large enough $n$ and $s = \Omega(\frac{n}{\epsilon^2})$, we conclude that:

$$\mathbb{P}\left(|\log\det \mathbf{Z}\mathbf{S}\mathbf{S}^T\mathbf{Z}^T - \log\det \mathbf{Z}\mathbf{Z}^T| < \epsilon\right) \geq 0.985. \tag{11}$$

We will use this inequality to bound the KL-divergence later. However, next, we bound the trace term. First, we rearrange the equation using the cyclic property of the trace:

$$\mathrm{tr}[(\mathbf{Z}\mathbf{Z}^T)^{-1}(\mathbf{Z}\mathbf{S}\mathbf{S}^T\mathbf{Z}^T)] = \mathrm{tr}[\mathbf{S}^T\mathbf{Z}^T(\mathbf{Z}^T\mathbf{Z})^{-1}\mathbf{Z}\mathbf{S}] = \mathrm{tr}[\mathbf{S}^T\mathbf{P}_\mathbf{Z}\mathbf{S}],$$

where $\mathbf{P}_\mathbf{Z} \in \mathbb{R}^{m\times m}$ is the projection to the row space of $\mathbf{Z}$. Note that the projection is idempotent, and so,

$$\mathrm{tr}[\mathbf{S}^T\mathbf{P}_\mathbf{Z}\mathbf{S}] = \mathrm{tr}[\mathbf{P}_\mathbf{Z}\mathbf{S}\mathbf{S}^T\mathbf{P}_\mathbf{Z}] = \mathrm{tr}\left[\mathbf{P}_\mathbf{Z}\cdot\mathbf{W}_m\left(s, \frac{1}{s}\cdot\mathbf{I}\right)\cdot\mathbf{P}_\mathbf{Z}\right],$$

where we use that $\mathbf{S}\mathbf{S}^T \sim \mathbf{W}_m(s, \frac{1}{s}\cdot\mathbf{I}_m)$, i.e., $\mathbf{S}\mathbf{S}^T$ is distributed as a $m\times m$ Wishart ensemble with $s$ degrees of freedom and covariance matrix equal to $\frac{1}{s}\cdot\mathbf{I}_m$. Since the Wishart ensemble is rotationally invariant (this immediately follows from rotational invariance of the multivariate Gaussian distribution), we can assume without loss of generality that $\mathbf{P}_\mathbf{Z}$ is the projection to the first $n$ standard basis vectors. From here it is easier to see that:

$$\mathrm{tr}\left[\mathbf{P}_\mathbf{Z}\cdot\mathbf{W}_m\left(s, \frac{1}{s}\cdot\mathbf{I}\right)\cdot\mathbf{P}_\mathbf{Z}\right] = \mathrm{tr}\left[\mathbf{W}_n\left(s, \frac{1}{s}\cdot\mathbf{I}\right)\right],$$

where we consider the size $n\times n$ Wishart matrix, since all entries not in the top $n\times n$ corner of $\mathbf{P}_\mathbf{Z}\cdot\mathbf{W}_m\left(s, \frac{1}{s}\cdot\mathbf{I}\right)\cdot\mathbf{P}_\mathbf{Z}$ are zero. We then observe that the trace of $\mathbf{W}_n\left(s, \frac{1}{s}\cdot\mathbf{I}\right)$ is the sum of the diagonal entries, and the $i$-th diagonal entry is equal to $\langle\mathbf{g}_i, \mathbf{g}_i\rangle$, where $\mathbf{g}_i \in \mathbb{R}^s$ and $\mathbf{g}_i \sim \mathcal{N}(\mathbf{0}, \frac{1}{s}\cdot\mathbf{I})$. Therefore,

$$\mathrm{tr}\left[\mathbf{W}_n\left(s, \frac{1}{s}\cdot\mathbf{I}\right)\right] = \sum_{i=1}^{n}\frac{1}{s}\cdot\sum_{j=1}^{s} g_{ij}^2,$$

where $g_{ij} \sim \mathcal{N}(0,1)$. Note that $g_{ij}^2$ is then a chi-squared random variable. The expectation of the above trace is $n$, and by Lemma 1 in [5],

$$\mathbb{P}\left(|\sum_{i=1}^{n}\frac{1}{s}\cdot\sum_{j=1}^{s} g_{ij}^2 - n| > \epsilon\right) \leq \mathbb{P}\left(|\sum_{i=1}^{n}\sum_{j=1}^{s} g_{ij}^2 - ns| > 4\sqrt{n}\cdot\frac{\epsilon\sqrt{s}}{4}\right) \leq \exp\left(\frac{-\epsilon^2 s}{16}\right).$$

Therefore, for every $\epsilon \in (0,1)$, we conclude the following:

$$\mathbb{P}\left(|\,\mathrm{tr}[(\mathbf{Z}\mathbf{Z}^T)^{-1}(\mathbf{Z}\mathbf{S}\mathbf{S}^T\mathbf{Z}^T)] - n| > \epsilon\right) \to 0 \quad \text{as } n\to\infty,\ \frac{s}{n} \geq 1. \tag{12}$$

Therefore, by eqns. (11, 12), for large enough $n$ and $s = \frac{Cn}{\epsilon^2}$, for some universal constant $C$, we have with probability at least 0.98 over the distribution of the Gaussian sketching matrix $\mathbf{S} \in \mathbb{R}^{m\times s}$ that,

$$\mathrm{D}_{\mathrm{KL}}(\mathcal{D}_2||\mathcal{D}_1) = \frac{1}{2}\left(\log\frac{\det \mathbf{Z}\mathbf{Z}^T}{\det \mathbf{Z}\mathbf{S}\mathbf{S}^T\mathbf{Z}^T} - n + \mathrm{tr}[(\mathbf{Z}\mathbf{Z}^T)^{-1}(\mathbf{Z}\mathbf{S}\mathbf{S}^T\mathbf{Z}^T)]\right) \leq \frac{1}{2}(\epsilon+\epsilon) \leq \epsilon.$$

Hence, we conclude the lemma statement.

We also note that $s \geq n$ is necessary for the statement to hold, otherwise the distributions would have different support and the KL-divergence would be infinite. This implies the dependency of our sketch size on $n$ in the above lemma is optimal.

We now show that one can perform a binary hypothesis test on the parameters of an LMM described in Section 2.1 by performing it instead on a sketched version of the model while only increasing the error probability of the test by an arbitrarily small term $\epsilon$. Intuitively, this means that we can use sketching to reduce the number of columns in the random effects matrix $\mathbf{Z}$ to a size that is linear in $n$ while preserving the information of the distribution.

**Theorem 3.** *Consider the binary hypothesis testing problem for the probabilistic model described in Section 2.1 with two sets of parameters,* $(\sigma_{g,0}^2, \tau_0)$ *and* $(\sigma_{g,1}^2, \tau_1)$*. Let* $\mathcal{D}_0 = \mathcal{N}(\mathbf{X}\beta, \sigma_{g,0}^2 \cdot \mathbf{Z}\mathbf{Z}^T + \tau_0 \mathbf{I}_n)$ *and* $\mathcal{D}_1 = \mathcal{N}(\mathbf{X}\beta, \sigma_{g,1}^2 \cdot \mathbf{Z}\mathbf{Z}^T + \tau_1 \mathbf{I})$ *represent the null and alternative hypothesis of the original model. Let* $\tilde{\mathcal{D}}_0 = \mathcal{N}(\mathbf{X}\beta, \sigma_{g,0}^2 \cdot \mathbf{Z}\mathbf{S}\mathbf{S}^T\mathbf{Z}^T + \tau_0 \mathbf{I}_n)$ *and* $\tilde{\mathcal{D}}_1 = \mathcal{N}(\mathbf{X}\beta, \sigma_{g,1}^2 \cdot \mathbf{Z}\mathbf{S}\mathbf{S}^T\mathbf{Z}^T + \tau_1 \mathbf{I})$*, where* $\mathbf{S} \in \mathbb{R}^{m \times k}$ *is a Gaussian sketching matrix, represent the corresponding sketched distributions.*

*If* $h : \mathbb{R}^n \to \{0, 1\}$ *is a hypothesis test which satisfies:*

$$\mathbb{P}_{\tilde{\mathcal{D}}_0}(h(\mathbf{y}) \neq 0) + \mathbb{P}_{\tilde{\mathcal{D}}_1}(h(\mathbf{y}) \neq 1) \leq \Delta,$$

*i.e., the sum of type one and type two error on the sketched testing problem is at most* $\Delta$*, then for* $s = \mathcal{O}(\frac{n}{\epsilon^4})$*, with probability at least* 0.98 *over the distribution of* $\mathbf{S}$*,*

$$\mathbb{P}_{\mathcal{D}_0}(h(\mathbf{y}) \neq 0) + \mathbb{P}_{\mathcal{D}_1}(h(\mathbf{y}) \neq 1) \leq \Delta + \epsilon.$$

*Proof.* First, by the data processing inequality (Theorem 1),

$$\mathrm{D}_{\mathrm{KL}}(\mathcal{D}_0 \;||\; \tilde{\mathcal{D}}_0) \leq \mathrm{D}_{\mathrm{KL}}(\mathcal{N}(\mathbf{0}, \mathbf{Z}\mathbf{Z}^T) \;||\; \mathrm{D}_{\mathrm{KL}}(\mathcal{N}(\mathbf{0}, \mathbf{Z}\mathbf{S}\mathbf{S}^T\mathbf{Z}^T)).$$

To see this, observe that given $\mathbf{r} \sim \mathcal{N}(\mathbf{0}, \mathbf{Z}\mathbf{Z}^T)$ and $\tilde{\mathbf{r}} \sim \mathcal{N}(\mathbf{0}, \mathbf{Z}\mathbf{S}\mathbf{S}^T\mathbf{Z}^T)$, we can sample $\mathbf{d} \sim \mathcal{N}(\mathbf{X}\beta, \tau_0 \mathbf{I})$. Then, $\sigma_{g,0} \cdot \mathbf{r} + \mathbf{d} \sim \mathcal{D}_0$ and $\sigma_{g,0} \cdot \tilde{\mathbf{r}} + \mathbf{d} \sim \tilde{\mathcal{D}}_0$. Notice that the randomness of $\mathbf{d}$ is independent from $\mathcal{D}_0$ and $\tilde{\mathcal{D}}_0$.

Next, let $\mathcal{H}_0$, $\tilde{\mathcal{H}}_0$, $\mathcal{H}_1$, and $\tilde{\mathcal{H}}_1$ denote the distribution of $h(\mathbf{y})$ where the distribution of $\mathbf{y}$ is given by $\mathcal{D}_0$, $\tilde{\mathcal{D}}_0$, $\mathcal{D}_1$, and $\tilde{\mathcal{D}}_1$ respectively. Any valid hypothesis test $h(\cdot)$ must be independent from $\mathcal{D}_0$ and $\tilde{\mathcal{D}}_0$, so we again apply the data processing inequality to conclude:

$$\mathrm{D}_{\mathrm{KL}}(\mathcal{H}_0 \;||\tilde{\mathcal{H}}_0) \leq \mathrm{D}_{\mathrm{KL}}(\mathcal{D}_0 \;||\; \tilde{\mathcal{D}}_0) \leq \mathrm{D}_{\mathrm{KL}}(\mathcal{N}(\mathbf{0}, \mathbf{Z}\mathbf{Z}^T) \;||\; \mathcal{N}(\mathbf{0}, \mathbf{Z}\mathbf{S}\mathbf{S}^T\mathbf{Z}^T)).$$

Hence, by Lemma 1 and Pinsker's inequality (Theorem 2),

$$\mathrm{D}_{\mathrm{TV}}\left(\mathcal{H}_0, \tilde{\mathcal{H}}_0\right) \leq \sqrt{\frac{1}{2}\mathrm{D}_{\mathrm{KL}}(\mathcal{N}(\mathbf{0}, \mathbf{Z}\mathbf{Z}^T) \;||\; \mathrm{D}_{\mathrm{KL}}(\mathcal{N}(\mathbf{0}, \mathbf{Z}\mathbf{S}\mathbf{S}^T\mathbf{Z}^T))} \leq \epsilon,$$

with probability at least 0.98 over the distribution of the Gaussian sketch $\mathbf{S}$ with $s = \mathcal{O}\left(\frac{n}{\epsilon^4}\right)$. By the same argument, $\mathrm{D}_{\mathrm{TV}}\left(\mathcal{D}_1, \tilde{\mathcal{D}}_1\right) \leq \epsilon$. Then,

$$|\mathbb{P}_{\mathcal{D}_0}(\mathbf{h}(\mathbf{y}) \neq 0) - \mathbb{P}_{\tilde{\mathcal{D}}_0}(\mathbf{h}(\mathbf{y}) \neq 0)| \leq \sup_{H \subset \{0,1\}} |\mathbb{P}_{\mathcal{H}_0}(\mathbf{h} \in H) - \mathbb{P}_{\tilde{\mathcal{H}}_0}(\mathbf{h} \in H)| = \mathrm{D}_{\mathrm{TV}}(\mathcal{H}_0, \tilde{\mathcal{H}}_0) \leq \epsilon.$$

By the same argument, $|\mathbb{P}_{\mathcal{D}_1}(\mathbf{h}(\mathbf{y}) \neq 1) - \mathbb{P}_{\mathcal{D}_1}(\mathbf{h}(\mathbf{y}) \neq 1)| \leq \epsilon$. We can now derive the statement of the theorem using the previous inequalities and the assumption on the hypothesis test $h(\cdot)$.

$$\mathbb{P}_{\tilde{\mathcal{D}}_0}(h(\mathbf{y}) \neq 0) + \mathbb{P}_{\tilde{\mathcal{D}}_1}(h(\mathbf{y}) \neq 1) \leq \Delta$$

$$\Rightarrow$$

$$\mathbb{P}_{\mathcal{D}_0}(h(\mathbf{y}) \neq 0) + \mathbb{P}_{\mathcal{D}_1}(h(\mathbf{y}) \neq 1)$$
$$\leq \Delta + |\mathbb{P}_{\mathcal{D}_0}(\mathbf{h}(\mathbf{y}) \neq 0) - \mathbb{P}_{\mathcal{D}_0}(\mathbf{h}(\mathbf{y}) \neq 0)| + |\mathbb{P}_{\mathcal{D}_1}(\mathbf{h}(\mathbf{y}) \neq 1) - \mathbb{P}_{\mathcal{D}_1}(\mathbf{h}(\mathbf{y}) \neq 1)|$$
$$\Rightarrow$$
$$\mathbb{P}_{\mathcal{D}_0}(h(\mathbf{y}) \neq 0) + \mathbb{P}_{\mathcal{D}_1}(h(\mathbf{y}) \neq 1) \leq \Delta + 2\epsilon.$$

Hence, we conclude the statement of the theorem after adjusting $\epsilon$ by a constant factor.

#### 1.B.1 Best Practices

The performance of our approach relies heavily on the sketching dimensions selected on the given dataset. If one is too stringent with the sketch dimension used for the samples, there may not be enough data present after sketching to yield accurate results. Similarly if the sketch dimension for the markers is too low, the resulting estimate for the relatedness matrix may not be accurate enough to yield meaningful results. However, if the sketch dimensions are too large (particularly the sample sketch dimension), that may result in diminishing returns. More specifically, the sketch dimension may be too large to experience improvements in execution time versus other state-of-the-art methods. Another bottleneck of a large sketching dimension is running into convergence issues when the dataset is too large after sketching.

The sketching parameters need to be tailored to each scenario to experience the best performance and results. For example, we would expect poor performance when applying MaSkLMM to a dataset with 10k samples and 100k markers using a sample sketch dimension of 5% as it will leave very little samples to perform accurately. However, if we apply the same sketch dimension to a dataset with 500k samples, then we would have plenty of information left after sketching to perform accurately. We see such tradeoffs in Figures 2 and 3.

#### 1.B.2 Quality Control

In this section, we discuss the parameters used for quality control (QC) and pruning in the real genotypes. Filtering was performed on both individuals and variants with at least 95% missing data. We checked for problematic sex assignment in missing gender fields using the X chromosome. We performed filtering for variants with minor allele frequency (MAF) $< 0.05$ and for variants which are not in Hardy-Weinberg equilibrium (HWE) with p-values at least 1e-16. This is done separately for cases and controls. We removed individuals with high or low heterozygosity rates and removed closely related individuals with Identity-by-descent (IBD) method owing to cryptic relatedness.

#### 1.B.3 UK Biobank Data

For the HYP and CAD dataset, the samples were extracted from UK Biobank data containing 331,256 European ancestry individuals and 411 parent phecode items (excluding infectious diseases, injuries, poisonings and pregnancy complications). This data was generated using a combination of NLP methods and manual curating to map ICD-10-CM codes to more meaningful phenotypes, clustered appropriately. More specifically, we included 6,300 ICD-10 diagnoses (data field 41270) with non-zero number of patients in our analyses. To reduce the dimensionality and increase the interpretability of our analyses, we further mapped the ICD-10 codes on to PheCode. Out of all 6,300 ICD-10 items, 4,807 could be mapped onto at least one valid PheCode, representing 505 PheCode and 1,434 child PheCode. We removed PheCode in categories that are dominated by non-genetic causes (infectious diseases, injuries & poisonings and pregnancy complications categories).

As a result of this process, we mapped a total of 4,004 ICD-10 codes to 411 parent PheCodes, including key ones such as hypertension and coronary artery disease, which were employed in our experiments.. [8]. Also, we downsampled the amount of controls in the data for CAD to experiment with varying ratios of cases to controls resulting in the 50k individuals.

#### 1.B.4 Additional Information

In this section, we assess the performance of MaSk-LMM through additional experiments on real and synthetic data.

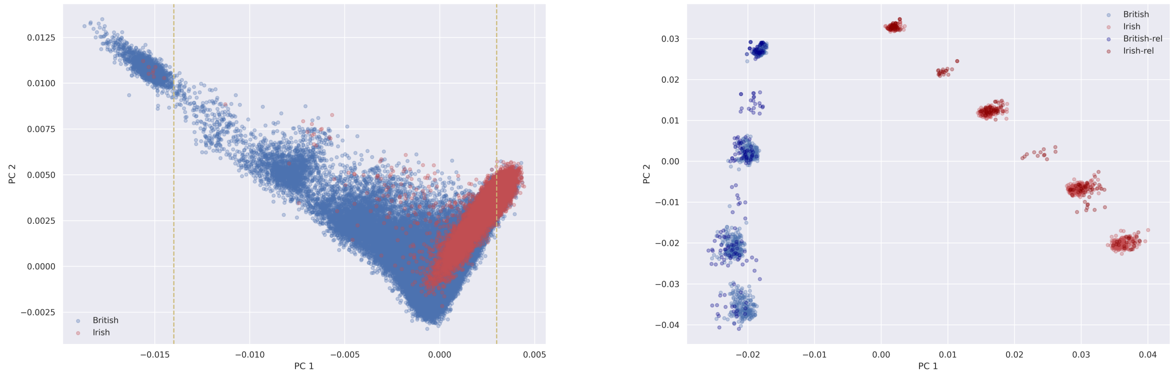


**(a)** **(b)**

**Fig. 1:** (a) Top two principal components of individuals with British and Irish ancestries from the UK Biobank data after performing quality control and pruning. The vertical lines show where we selected British and Irish samples to use in our simulation. (b) Using that subset as ancestors in the "mosaic-chromosome" scheme, we generated 10k synthetic individuals and plotted the top two principal components.

**Table 1:** Average execution time (in minutes) and causal/spurious associations captured by MaSk-LMM (varied sample sketch dimension), Regenie, and BOLT-LMM when applied to the simulated dataset, $D_1$. The minimum and maximum times are shown in parentheses.

| | MaSk (0.1) | MaSk (0.2) | MaSk (0.3) | Regenie | BOLT |
|---|---|---|---|---|---|
| time | 0.5 (0.4/0.5) | 0.7 (0.5/0.7) | 1.0 (0.6/1.1) | 30.5 (26.5/35.2) | 22.0 (18.7/25.1) |
| assocs. | 4 (0) | 8 (1) | 13 (2) | 13 (3) | 13 (3) |

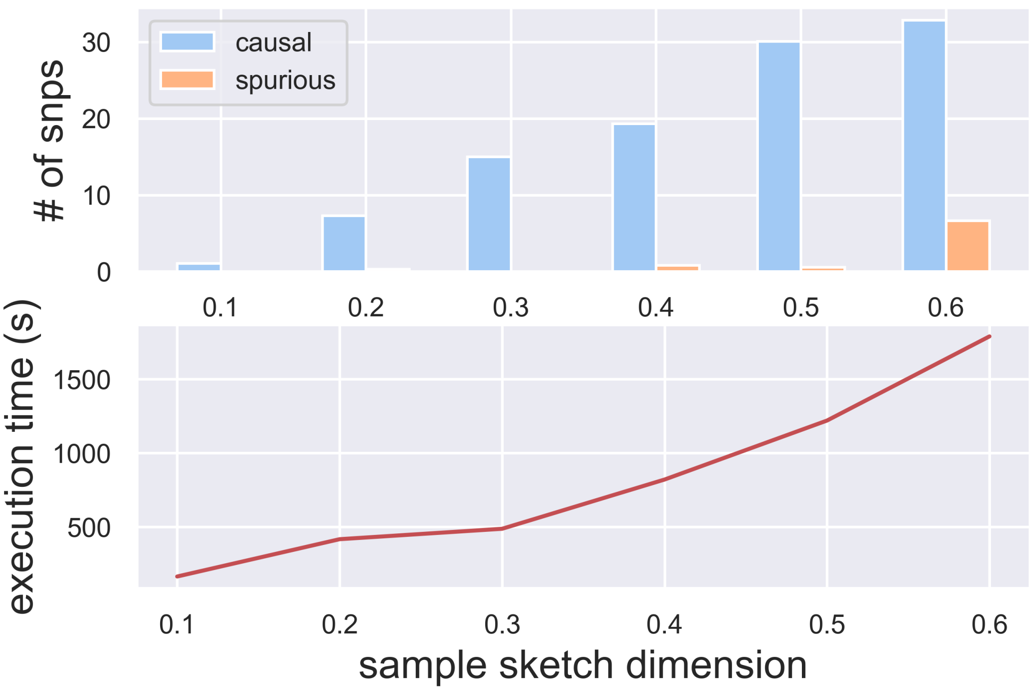


**Fig. 2:** Performance of MaSk-LMM as we vary the sketching dimension used on the number of samples. We applied MaSk-LMM on a simulated dataset of 10k individuals and approximately 250k SNPs with about 100 SNPs set as causal using no sketching for the markers. We repeated this evaluation 10 times and report the average number of causal associations, spurious associations and execution time.

**Table 2:** Average execution time (in minutes) and causal/spurious associations captured by MaSk-LMM (varied sample sketch dimension), Regenie, and BOLT-LMM when applied to the simulated dataset, $D_2$. The minimum and maximum times are shown in parentheses.

| | MaSk (0.1) | MaSk (0.2) | MaSk (0.3) | Regenie | BOLT |
|---|---|---|---|---|---|
| time | 15.2 (14.6/15.8) | 74.0 (68.4/79.7) | 197.3 (167.6/232.9) | 309.3 (289.6/335.7) | 219.6 (209.5/231.8) |
| assocs. | 16 (4) | 18 (20) | 19 (28) | 20 (45) | 7 (35) |

**Table 3:** Average execution time (in minutes) and causal/spurious associations captured by MaSk-LMM (varied sample sketch dimension), Regenie, and BOLT-LMM when applied to the simulated dataset, $D_3$. The minimum and maximum times are shown in parentheses.

| | MaSk (0.03) | MaSk (0.04) | MaSk (0.05) | Regenie | BOLT |
|---|---|---|---|---|---|
| time | 37.1 (35.1/40.1) | 56.8 (55.2/61.5) | 88.5 (85.0/94.7) | 911.4 (866.5/950.7) | 1674.5 (1643.9/1747.1) |
| assocs. | 13 (2) | 16 (9) | 21 (34) | 24 (220) | 9 (130) |

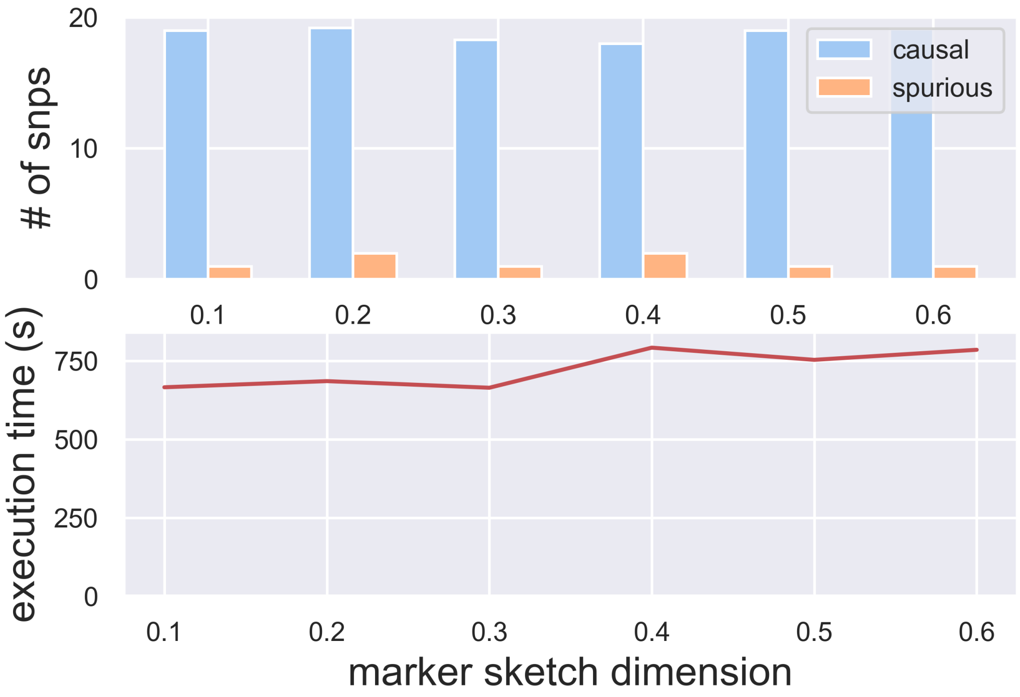


**Fig. 3:** Performance of MaSk-LMM as we vary the sketching dimension used on the number of markers when computing the GRM. We applied MaSk-LMM on a simulated dataset of 10k individuals and approximately 250k SNPs with about 100 SNPs set as causal using a constant sketch dimension of 0.4 for the samples. We repeated this evaluation 10 times and report the average number of causal associations, spurious associations and execution time.

**Table 4:** Number of causal associations and execution time of MaSk-LMM applied to $D_1$ (British-Irish data with 10k samples and 265k SNPs) and varying the sample sketch dimension with no marker sketching.

| Sketching dimension | 0.1 | 0.2 | 0.3 | 0.4 | 0.5 | 0.6 | 0.7 | 0.8 | 0.9 | 1.0 |
|---|---|---|---|---|---|---|---|---|---|---|
| Num. causal | 5 | 14 | 15 | 16 | 16 | 15 | 17 | 17 | 18 | 21 |
| Execution time (hours) | 0.025 | 0.03 | 0.05 | 0.1 | 0.2 | 0.3 | 0.5 | 0.7 | 0.9 | 1.2 |

**Table 5:** Number of causal associations and execution time of MaSk-LMM applied to $D_1$ (British-Irish data with 10k samples and 265k SNPs) and varying the marker sketch dimension with no sample sketching. When setting marker sketching to 10%, the method did not converge (entry set to "n/a").

| Sketching dimension | 0.1 | 0.2 | 0.3 | 0.4 | 0.5 | 0.6 | 0.7 | 0.8 | 0.9 | 1.0 |
|---|---|---|---|---|---|---|---|---|---|---|
| Num. causal | n/a | 15 | 16 | 17 | 17 | 17 | 17 | 17 | 17 | 17 |
| Execution time (hours) | 1.7 | 1.3 | 1.1 | 0.9 | 0.9 | 1.1 | 1.1 | 1.2 | 1.2 | 1.1 |

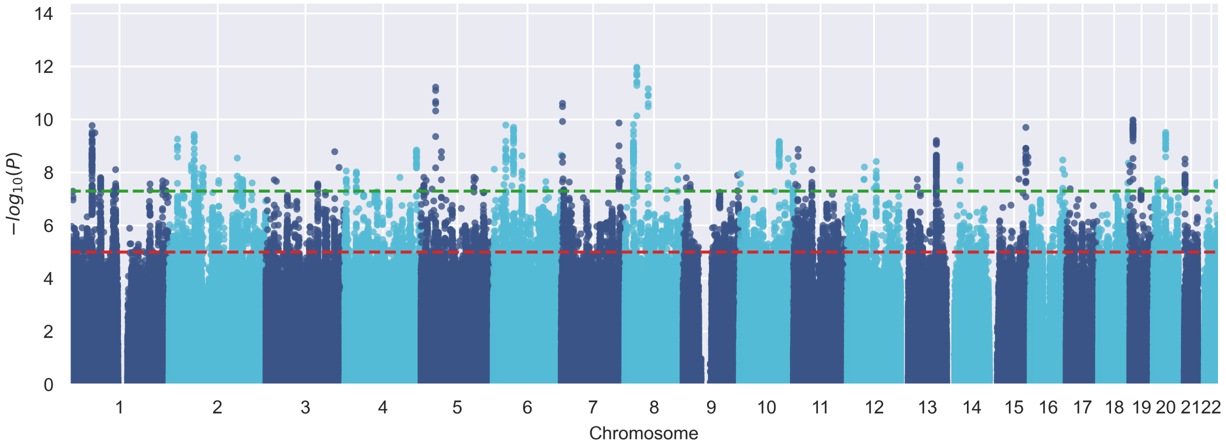


**Fig. 4:** Manhattan plot for Hypertension.

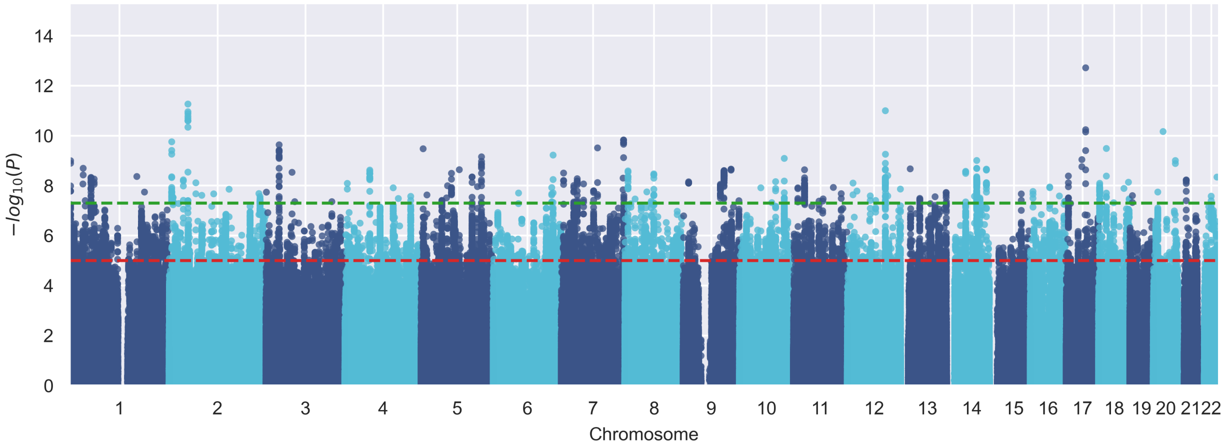


**Fig. 5:** Manhattan plots for Coronary Artery Disease.
